# RT2C: Predicting Time to New Dental Caries Using a Recurrent Neural Network Model Trained on Multi-Site Longitudinal Structured Dental Data

**DOI:** 10.1101/2025.09.03.25334405

**Authors:** Xiangyi Liu, Krishna Kumar Kookal, Ryan Brandon, D. Brad Rindal, Joanna Mullins, Joel M. White, Muhammad F. Walji, Laila Rasmy

## Abstract

Dental caries remains one of the most prevalent chronic conditions globally, yet efforts to quantify a patient’s risk of developing new caries over time remain limited. Existing caries risk assessment (CRA) tools are widely used in clinical settings but typically rely on cross-sectional data, which may limit their predictive accuracy and generalizability across populations. To address these limitations, we developed RT2C, a recurrent neural network model based on bidirectional gated recurrent units (Bi-GRU) to predict time to new caries. The model was trained on longitudinal dental records from 466,782 patients across four diverse dental organizations, encompassing over one million visits between 2019 and 2023. Input features included demographics, preventive procedures, caries history, and previously recorded CRA scores. We benchmarked the model performance against other machine learning models such as logistic regression, light gradient boost machine, random survival forest, and CRA-based baselines using AUROC for binary classification and concordance index (c-index) for survival prediction.

Our model demonstrated strong predictive performance, achieving a c-index of 88.52% (95% CI: 88.51-88.54) in survival analysis, significantly outperforming baseline models based on current or past CRA scores by more than 18%. Site-level evaluations confirmed its robust generalizability, with pooled models performing comparably or better than sites-pecific ones. Additionally, decision curve analysis showed that the Bi-GRU model offers greater clinical net benefit across a range of decision thresholds when compared to CRA or treat-all/treat-none strategies. By leveraging longitudinal dental histories and temporal patterns in patient visits, our proposed model provides clinically meaningful improvements in predicting caries risk. These findings support its potential integration into decision support tools for more personalized and preventive dental care.

## Introduction

According to Global Burden of Disease Study 2021, untreated dental caries of permanent teeth had a global age-standardized prevalence of 27.5% in 2019, making it one of the most common chronic conditions worldwide. Although the age-standardized prevalence of caries in permanent and deciduous teeth decreased slightly by 3.6% and 3.0%, respectively over the past decade (Wen et al., 2021), untreated caries remains a major public health concern. Multiple factors contribute to an individual’s caries risk, including past caries experience, poor oral hygiene, enamel defects, and dietary behaviors such as high sugar intake (Ng et al. 2024).

Several caries risk assessment (CRA) tools have been developed (Bratthall and Petersson, 2005; Featherstone and Chaffee, 2018; Gao et al. 2010; Brons-Piche et al. 2019; Paul et al. 2025; Lucchi et al. 2024). These tools differ in both the domains of information they consider and the methodological approaches they employ for risk estimation, and have demonstrated generally acceptable performance in binary prediction of future caries, with reported area under the receiver operating characteristic curve (AUROC) ranging from modest to good across populations (Su et al. 2021). However, evidence from systematic reviews indicates that most existing validation studies carry a high risk of bias, largely due to insufficient sample sizes, substantial loss to follow-up, and suboptimal handling of missing data (Su et al. 2021). Therefore, there is a need to enhance existing CRA tools by integrating advanced analytical methods with large-scale EHR data, enabling more precise and clinically relevant predictions of caries outcomes.

Accurately identifying which patients are most likely to develop new caries is critical for enabling dentists to provide appropriate preventive care, tailor recall intervals, and optimize resource allocation. Recent advances in dental informatics have enabled the development of new outcome measures using electronic health record (EHR) data to track the occurrence of new tooth decay (Brandon et al. 2022), creating opportunities for continuous risk monitoring and quality improvement. Capitalizing on these opportunities requires predictive models that can fully exploit the longitudinal and updatable nature of EHRs. However, many current tools and models fall short, as they are constrained by smaller cohorts used and are not designed to sufficiently leverage sequential visit information from patient’s history (Wang et al. 2025; Ramos-Gomez et al. 2021; Sadegh-Zadeh et al. 2022; Park et al. 2021; Kang and Njimbouom et al. 2022).Furthermore, most current machine learning models focus on binary prediction, which requires setting probability thresholds. This approach can result in information loss and potential bias, whereas incorporating time-to-event prediction with continuous risk estimation can provide more nuanced and clinically actionable assessments (Moons et al. 2019).

To address these limitations, we developed RT2C (Recurrent neural network-based model for Time to Caries), a generalizable RNN-based approach that integrates multi-domain features from patients’ longitudinal dental records and leverages temporal patterns across visits to estimate continuous caries risk. Importantly, our model aims to move beyond binary classification by predicting continuous risk scores for new caries at subsequent visits, which commonly occur at variable time intervals, thereby enhancing the precision and clinical relevance of caries risk assessment. A key strength of our study besides the proposed model architecture lies in the large, diverse dataset, which includes over 460,000 patients spanning all age groups and drawn from four distinct dental care organizations. This multi-site, heterogeneous sample enhances the model’s generalizability and provides a more comprehensive representation of real-world dental populations, addressing common limitations in prior studies that relied on smaller or more narrowly defined cohorts.

## Methods

This study utilized retrospective, patient-level data collected from four participating dental care organizations: two dental schools, one dental accountable care organization (ACO), and one integrated healthcare system offering dental plans, including HMO/DMO and PPO options. Data from dental schools and the ACO were previously used in a related study on tooth decay quality measures (Brandon et al., 2022), while the DMO was added as an additional site under the current study. The usage of limited/deidentified data derived from electronic health records (EHR) of the four sites for secondary analysis was approved by the Institutional Review Board (IRB) under the approval number: HSC-DB-21-0471.

Although the DMO site used the Epic EHR platform (Epic Systems, Verona, WI), it implemented scripts to achieve consistent data elements as the other three sites used the axiUm EHR platform (Henry Schein Corp.). Furthermore, while the DMO site utilized the systematized nomenclature of dentistry (SNO-DENT), and the other three sites used the systematized nomenclature dental diagnostic system (SNO-DDS) dental diagnostic terminology, we completed testing and validation to show consistent results in the data collection and outcome calculation process. All four sites have established comprehensive training programs, standardized practice guidelines, clinical decision support tools, and quality assurance systems. Data were extracted from the EHR of four sites spanning five calendar years, from January 1, 2019, to December 31, 2023, including patients covering all age groups.

The model was trained and validated on the oral health data containing visit-level relevant dental data contributed by 4 different sites for a total of 466,782 patients (Table 1). 413,864 visits were excluded because they lacked both prior visits and the caries outcome at the visit for model training or testing. The model was trained on heterogeneous data of 326,748 patients (744,624 visits), validated on data of 46,678 patients (106,616 visits), and tested on data of 93,356 patients (211,313 visits). There was no overlapping on the patient level across training, validation, and test sets.

**Table 1.** Descriptive statistics of study population based on visit-level counts.

|  | Site 1 | Site 2 | Site 3 | Site 4 | Total |
| --- | --- | --- | --- | --- | --- |
| Number of Patients, n | 18,002 | 22,555 | 334,628 | 91,597 | 466,782 |
| Number of Visits, n | 43,194 | 49,473 | 703,726 | 266,160 | 1,062,553 |
| Gender, n (%) |  |  |  |  |  |
| Female | 23,430 (54) | 24,843 (50) | 400,368 (57) | 154,650 (58) | 603,291 (57) |
| Male | 19,689 (46) | 24,609 (50) | 301,970 (43) | 111,493 (42) | 457,761 (43) |
| Other | 75 (0) | 21 (0) | 1,388 (0) | 17 (0) | 1,501 (0) |
| Race, n (%) |  |  |  |  |  |
| Caucasian | 11,781 (27) | 9,119 (18) | 440,847 (63) | 175,349 (66) | 637,096 (60) |
| African American | 6,089 (14) | 2,982 (6) | 18,947 (3) | 32,326 (12) | 60,344 (6) |
| Asian | 2,734 (6) | 8,682 (18) | 34,087 (5) | 20,856 (8) | 66,359 (6) |
| Other | 2,951 (7) | 5,258 (11) | 110,777 (16) | 11,116 (4) | 130,102 (12) |
| Unknown | 19,639 (45) | 23,432 (47) | 99,068 (14) | 26,513 (10) | 168,652 (16) |
| Age (years), n(%) |  |  |  |  |  |
| 0 ~ 5 | 7,045 (16) | 6,511 (13) | 27,628 (4) | 11,629 (4) | 52,813 (5) |
| 6 ~13 | 10,314 (24) | 13,903 (28) | 110,708 (16) | 45,508 (17) | 180,433 (17) |
| 14~16 | 3,302 (8) | 2,378 (5) | 45,049 (6) | 12,835 (5) | 63,564 (6) |
| 17 and above | 22,533 (52) | 26,681 (54) | 520,341 (74) | 196,188 (74) | 765,743 (72) |
| CRA, n(%) |  |  |  |  |  |
| Low | 2,479 (6) | 3,197 (6) | 411,058 (58) | 109,799 (41) | 526,533 (50) |
| Moderate | 7,938 (18) | 9,340 (19) | 155,113 (22) | 87,440 (33) | 259,831 (24) |
| High or Extreme | 23,721 (55) | 23,781 (49) | 136,879 (19) | 57,375 (22) | 241,756 (23) |
| Unknown | 9,056 (21) | 13155 (27) | 676 (0) | 11,546 (4) | 34,433 (3) |
| Insurance, n(%) |  |  |  |  |  |
| Private/Third-party | 8,062 (19) | 15,540 (31) | 615,502 (87) | 201,158 (76) | 840,262 (79) |
| Government | 16,767 (39) | 26,983 (55) | 81,687 (12) | 64,368 (24) | 189,805 (18) |
| Cash/No Insurance | 18,365 (42) | 6,950 (14) | 6,537 (1) | 634 (0) | 32,486 (3) |
| Duration between visits (months) |  |  |  |  |  |
| Median (IQR) | 9 (7-13) | 12 (8-17) | 16 (13-21) | 11 (8-16) | 15 (12-19) |
| Duration between the index visit<sup>1</sup> and the next visit with new caries (days) |  |  |  |  |  |
| Median (IQR) | 11 (7-17) | 15 (10-24) | 17 (13-24) | 15 (10-22) | 16 (12-23) |

1. Index visit: A visit that serves as the prediction timepoint.

The dataset used in this study was generated through a Common Data Model (CDM) designed to enable structured, visit-level data extraction from electronic health records (EHRs) across multiple sites. A structured query language (SQL) script was developed to retrieve relevant documentation from consistent locations within the axiUm EHR system, initially deployed at three participating sites. To ensure consistency across sites, the extracted data were mapped to a standardized format of diagnoses and procedures. For sites using the Epic EHR system, equivalent SAS scripts were developed after assessing data availability and compatibility for each quality measure. The process was iteratively assessed by clinical experts and informaticists. The CDM was specifically designed to comprehensively capture all data elements necessary for evaluating established dental quality measures, including those related to caries risk. Prior work has demonstrated the feasibility and validity of using EHR-based measures to assess caries risk and related preventive services across multiple settings (Kumar et al. 2018; Bangar et al. 2022; Neumann et al. 2019). Based on this foundation, our study utilized all features available in the aforementioned CDM and had previously shown to be relevant and valid for assessing caries risk in pediatric and general dental populations.

Our model input variables included both continuous and categorical features. The categorical features include sites, demographics (gender, race/ethnicity), insurance type (cash, government, private, third-party), Caries Management by Risk Assessment (CAMBRA®)-caries risk assessment (CRA) at the current visit, and CAMBRA-CRA at the previous visit. The continuous features include age at the time of visit, the number of dental visits in the past year, the number of teeth eligible for sealants, the number of sealed teeth, and the number of days past since the most recent preventive measures (fluoride procedure, silver diamine fluoride procedure, high fluoride toothpaste prescription, and chlorhexidine prescription). Based on our initial experiments, we found that when continuous features were treated as categorical and embedded along with other categorical features was a higher performance gain (Appendix Table 1). Therefore, we converted our continuous variables into categorical variables by assigning each unique value a distinct category.

As illustrated in Fig. 1, we defined index visit (Visit i, 2 ≤ i < k) as the visit that the model predicts the risk for dental caries to be diagnosed by the next visit, using the history of all previous visits and the measurements at the index visit. Although the predicted visit data used in our study is derived from EHR data, it shares similarities with structured EHR. We thereby inherited the classical formulation of EHR from literature, where the dental history was as a list of visits, and each visit as a list of codes representing the records measured at the visit (Rasmy et al. 2021). For a detailed description of the data structuring and model architectures, please refer to the Supplementary Appendix.

**Fig 1.**
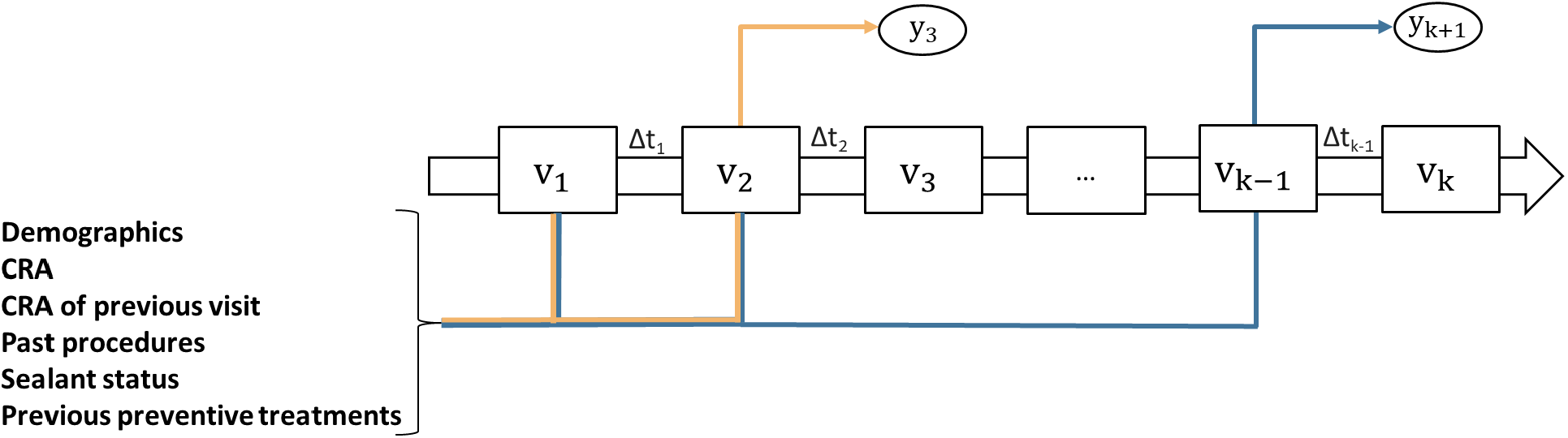
Prediction tasks: To predict the occurrence of new caries at Visit k, the dental history from all previous visits is consumed by the proposed RNN model. V_i_ represents the i^th^ visit, which includes patient demographics, CRA, CRA of the previous visit, number of procedures received in the past 3 years, sealant status, previous preventive treatments (e.g., fluoride procedure, silver diamine fluoride procedure, high-fluoride toothpaste prescription, chlorhexidine prescription), and the duration between visits (Δt_i_). At each step, the model uses dental history up to Visit i to generate a risk prediction y_i+1_ for the subsequent visit (i+1), thereby leveraging sequential and temporal patterns across the longitudinal dental record.

RNN models have demonstrated sufficient predictive power for clinical event prediction using EHR data, while remaining relatively simple in architecture (Rasmy et al. 2021). Given that our data is sequential and it uses longitudinal data from previous visits, we proposed a bidirectional RNN-based model, R2TC, which utilizes gated recurrent units (Bi-GRU). In this architecture, the model processes the patient’s history until the visit where the prediction is conducted in both forward and backward directions, allowing it to learn patterns that depend on the order and timing of visits (Jagannatha and Yu, 2016). Inherited from the Pytorch_EHR framework (source code: https://github.com/ZhiGroup/pytorch_ehr), the model was designed to consume the entire patient history from EHR without requiring special formatting for different data types or imputation for missing values. This approach enables the model to take advantage of all available information in a patient’s history and has been shown to perform well for predicting multiple clinical outcomes (Rasmy et al. 2022; Nigo et al. 2024). For the binary prediction task, we compared our proposed model to two traditional machine learning algorithms: logistic regression (LR) and light gradient boosting machine (LGBM). For the survival prediction task, we compared the predictive performance of our model against random survival forest (RSF) and two baselines: one based solely on the CAMBRA-CRA evaluation at the current visit, and another using the most recent CAMBRA-CRA evaluation from a prior visit. Given its widespread use in clinical practice, CAMBRA-CRA serves as a meaningful benchmark. It is a standardized clinical tool for assessing caries risk, incorporating biological, behavioral, and clinical indicators to assign patients to risk categories. It has been routinely used in practice since 2003 and has shown strong predictive validity for future caries development in longitudinal studies (Featherstone and Chaffee, 2018; White et al., 2020). To enable comparison with CRA-based risk groupings, we defined model-predicted risk categories based on reference percentiles from the validation set: patients were classified as low risk if their predicted risk score fell below the 20th percentile, moderate risk if between the 20th and 70th percentiles, and high risk if above the 70th percentile.

The primary outcome of our study was the risk of developing new caries at the next dental visit. In the binary prediction task, this outcome was represented as a binary indicator of whether new caries occurred at the next visit. In the survival prediction task, it was modeled as the time-to-event for new caries at the next visit. To assess the discriminative ability of the proposed model, we used the AUROC as the model performance metric for the binary classification model. We also reported the area under the precision-recall curve (AUPRC) and clinically relevant metrics: specificity at 95% sensitivity, F1-score, and the sensitivity and specificity at the optimum threshold (the cutoff probability that leads to the highest sensitivity and specificity). For the survival prediction task, we reported the concordance index (c-index) as our primary metric. To evaluate the clinical utility of our predictive model, we performed a decision curve analysis (DCA). DCA allows for direct assessment of net clinical benefit across a range of decision thresholds, offering insight into the value of model-guided decisions in real-world settings (Vickers et al. 2019).

## Results

The study was conducted on the cohort of patients who, on average, were 36.5 years old (std: 22.5; median: 36; interquartile range: 15-55) and their 1,062,553 dental visits. Each patient had, on average, 2.28 (std: 1.46) visits. The mean days difference between visits is 311.4 (std: 169.8). For the survival prediction task, our proposed model achieves a c-index of 88.52% (95% CI: 88.51-88.54%). It outperformed the RSF by 6% (c-index: 81.60%, 95% CI: 81.48-81.72%). To isolate the impact of longitudinal data on model’s predictive power, we compared the model performance using the entire visit history (c-index: 88.52%, 95% CI: 88.51-88.54%) to the model using only the latest visit (c-index: 85.42%, 95% CI: 85.40-85.44%), with the difference being significant (p-value < 0.005). RNN models with other types of cell units are detailed in Appendix Table 2. The prediction performance of the proposed RT2C model was evaluated across varying durations between visits, as detailed in Appendix Table 3.

To assess the generalizability of our model, we compared the performance of pooled and site-specific models on each site’s test dataset using the concordance index (c-index) for the survival prediction task. In three out of four sites, the pooled Bi-GRU model achieved superior or comparable performance relative to models trained exclusively on local site data (Table 2). Only one site demonstrated improved performance with its site-specific model, indicating that the pooled model maintains robust generalizability across diverse institutional settings while minimizing the need for local retraining.

**Table 2.** Comparison of survival prediction performance between site-specific models and the proposed general model.

| Test c-index (std) | Combined | Site 1 | Site 2 | Site 3 | Site 4 |
| --- | --- | --- | --- | --- | --- |
| RT2C (General Bi-GRU model) | **88.2** (0.47) | <u>78.4</u> (0.47) | **80.9** (0.48) | **87.3** (0.78) | **86.9** (0.28) |
| Site-specific Bi-GRU model | N/A<sup>1</sup> | **80.2** (0.95) | <u>80.0</u> (0.36) | <u>86.0</u> (0.17) | <u>86.6</u> (0.25) |
| RandomSurvivalForest | 81.6 (0.03) | 66.5 (0.89) | 68.6 (0.71) | 81.7 (0.14) | 73.2 (0.08) |
| Baseline Model 1 - Predict on current CRA | 70.2 (0.10) | 57.6 (0.75) | 58.1 (0.36) | 69.5 (0.17) | 67.1 (0.12) |
| Baseline Model 2 - Predict on last CRA | 65.2 (0.13) | 57.6 (0.39) | 57.7 (0.45) | 61.5 (0.13) | 63.9 (0.37) |

<sup>1</sup>Site-specific models are not tested on the combined test set.
The numbers in boldface indicate the highest c-index per task; the numbers underlined represent the second highest.

To assess the survival prediction performance of our model, we compared the predictive ability of our model against two baseline models that used only the CAMBRA-CRA measures, which are widely used in routine clinical practice to estimate a patient’s caries risk (Featherstone and Chaffee, 2018). When applied to the same test cohort, the baseline model using CRA assessed at the current visit achieved a c-index of 70.2%, while performance declined further to 65.2% when using CRA from the most recent prior visit. In contrast, our proposed Bi-GRU model, which integrates longitudinal and multi-domain features, consistently outperformed both baselines by a margin of at least 18 percentage points in c-index. Site-level analyses further confirmed the superior performance of our model relative to CRA-only approaches, both at the current and past visit levels (Table 2), reinforcing its potential to enhance caries risk prediction across diverse clinical settings. To further evaluate the clinical utility of our model, we examined cases where CAMBRA-CRA classifications appeared misaligned with actual caries outcomes. As shown in Table 3, a substantial proportion of false negatives (FNs)—patients incorrectly labeled as low risk by CAMBRA-CRA but who later developed new caries—were successfully identified as high risk by RT2C. Across sites, RT2C correctly flagged between 73.65% and 96.03% of these CRA FNs. Conversely, among false positives (FPs)—patients labeled as high/extreme risk by CRA but who did not develop new caries—RT2C identified a smaller but non-negligible proportion as low risk, with corrective rates ranging from 1.23% to 26.19%.

**Table 3.** Percentage of CAMBRA-CRA misclassifications corrected by RT2C across test sites.

|  | Site 1 | Site 2 | Site 3 | Site 4 |
| --- | --- | --- | --- | --- |
| % CRA False Negatives Corrected by RT2C (std) | 96.03 (0.69) | 73.65 (2.91) | 76.52 (0.94) | 88.99 (2.1) |
| % CRA False Positives Corrected by RT2C (std) | 2.82 (0.94) | 6.23 (1.87) | 26.19 (6.39) | 1.23 (1.70) |

We evaluated the model’s discriminative ability across various patient subgroups defined by insurance type, age group, gender, CRA level, and CRA level measured at the last visits (CRA_Past). The c-index values consistently exceeded 0.81 across all subgroups, indicating robust predictive performance. Among insurance categories, the model achieved the highest performance for patients with private/third-party insurance (c-index = 0.8857), followed closely by government-covered patients, while slightly lower for patients paying with cash or without insurance (c-index = 0.8120). Age-based performance was highest for adolescents aged 14–16 (c-index = 0.9011), and lowest among children aged 0–5 (c-index = 0.8704). Gender-based results were comparably strong across all groups, with c-index values ranging from 0.8802 to 0.8966. For racial subgroups, performance was strongest in White patients (c-index = 0.8871), followed by Other (c-index = 0.8730) and Black patients (c-index = 0.8668). Performance also improved with increasing CRA levels and CRA_Past levels, with the greatest performance observed in patients with “High and Extreme” risk (c-index = 0.8604 for CRA and 0.8745 for CRA_Past).

Additionally, our DCA, as shown in Figure 4, shows that our proposed model achieved the highest net benefit across most threshold probabilities compared to both CRA and CRA Past. Specifically, it maintained a positive net benefit from threshold probabilities of 0.1 to 0.7, where CRA-based tools either declined or showed minimal added benefit. This indicates that the model may lead to better clinical outcomes by guiding more efficient and targeted preventive strategies.

## Discussion

We trained a dental caries risk predictive model on data from over 460,000 patients across all age groups and more than one million dental visits. Our model outperformed the current caries risk assessment tool in use (CAMBRA-CRA), which has demonstrated clear connections between its risk levels and the occurrence of new caries (White et al. 2020). Our proposed model R2TC took into consideration the time to new caries, and it showed a higher concordance index (c-index > 0.88) vs CAMBRA-CRA (c-index = 0.70) for identifying new caries at the next dental visit. Additionally, our deep learning based algorithm outperformed the machine learning based RSF model (c-index = 0.82). Furthermore, our proposed model exhibited robust generalizability across different sites (Table 2) and patient subgroups based on demographic and socioeconomic factors (Fig. 3). The proposed model also demonstrated clearer separation between risk groups, particularly between moderate and high risk during early follow-up, indicating improved early detection of elevated caries risk (Fig. 2). It outperformed CAMBRA-CRA by leveraging longitudinal, multi-domain data to identify high-risk individuals often misclassified as low risk by CRA. Notably, among those mislabeled as low risk by CRA but who developed new caries, 70–95% were correctly flagged by our RT2C model with event probabilities above 0.2 across sites (Table 3). This indicates the model’s strong ability to recover high-risk individuals missed by traditional clinical risk assessments. Conversely, RT2C also correctly reclassified a proportion of patients labeled high risk by CRA but who did not experience caries, assigning them low predicted probabilities. While the magnitude of this correction varied by site, it demonstrates RT2C’s added value in both improving sensitivity and reducing overtreatment based on misclassified CRA scores. Additionally, the decision curve analysis (DCA) shows that our model consistently offers greater net clinical benefit than CRA-based baselines across a wide range of threshold probabilities (0.1–0.7). This suggests that the model supports more effective risk-based decision-making by identifying more true positives while minimizing unnecessary interventions. Its sustained net benefit across thresholds highlights its potential for flexible, actionable use in preventive care planning.

**Fig 2.**
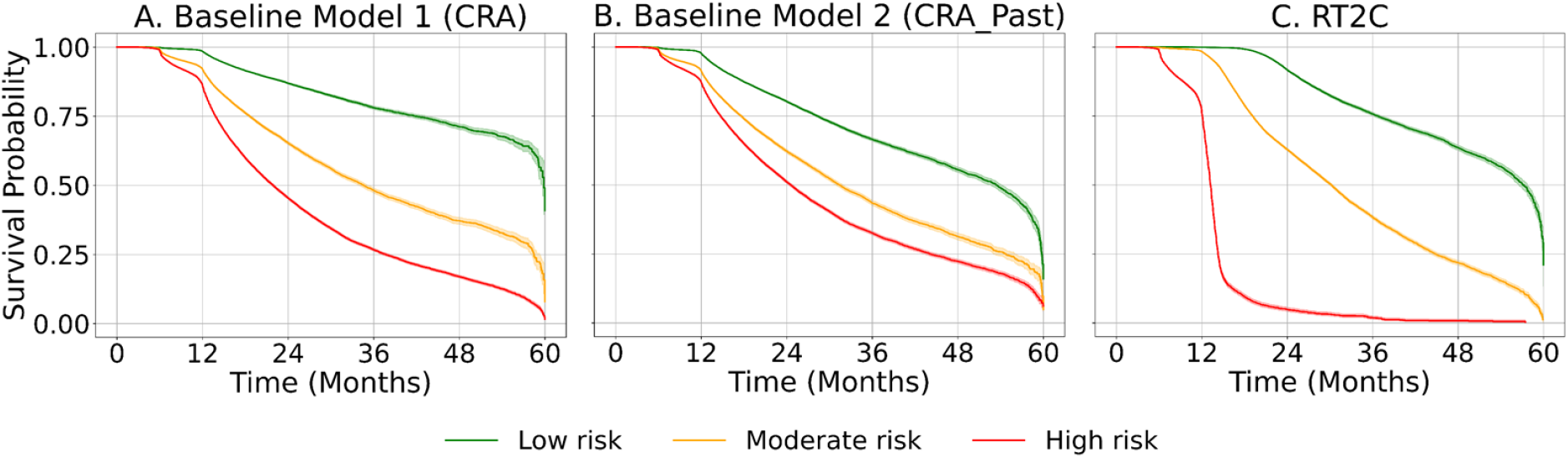
Survival curves are shown for the baseline model using current CRA levels (A), baseline model using CRA from prior visits (B), and the proposed RT2C model (C). Risk groups for RT2C are defined based on predicted risk percentiles from the validation set: ‘low risk’ (<20th percentile), ‘moderate risk’ (20th–70th percentile), and ‘high risk’ (>70th percentile). Patients are stratified by their respective risk group, and curves represent predicted time to new caries occurrence at each patient visit level.

**Fig 3.**
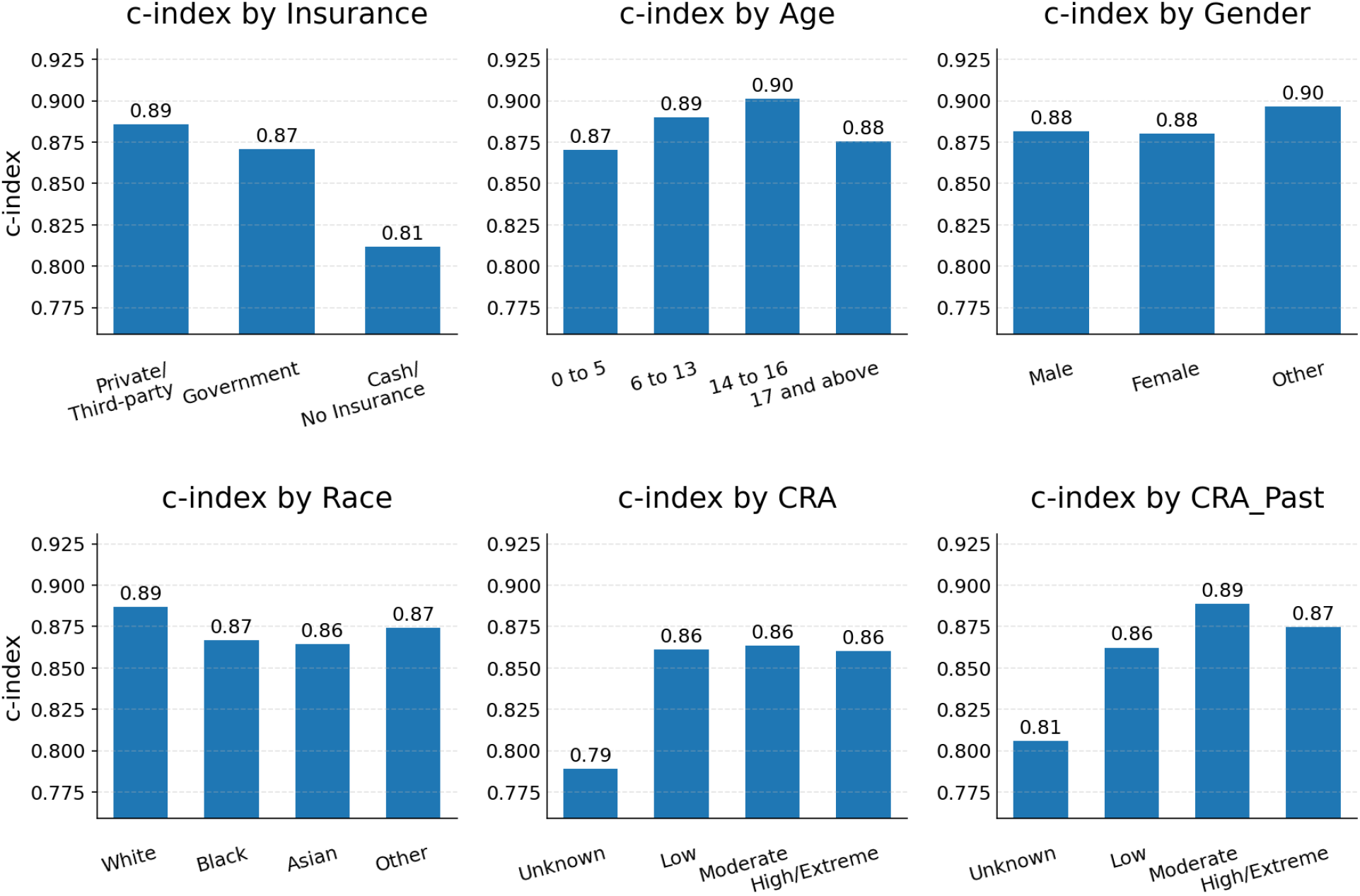
Subgroup analysis by insurance types, age groups, gender, CRA, and CRA measured at the previous visits (CRA_Past).

**Fig 4.**
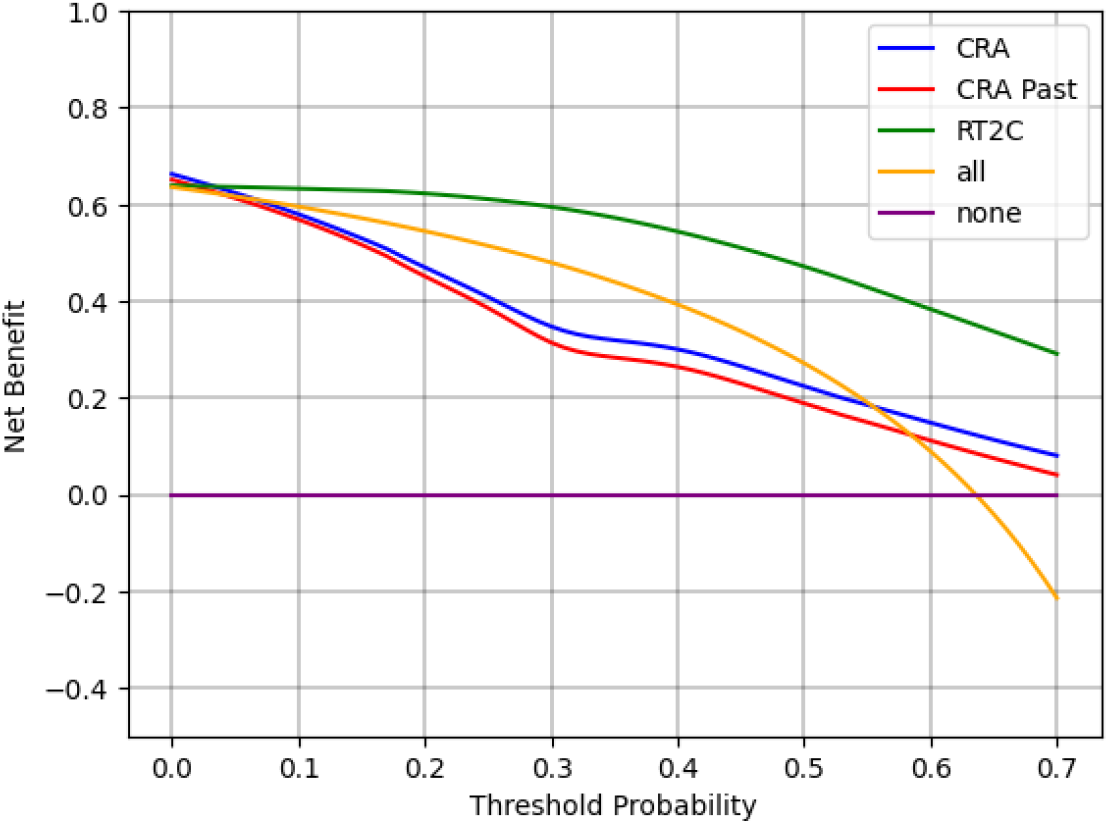
Decision Curve Analysis comparing net benefit across different threshold probabilities for the proposed model (green), CRA-based risk (blue), and prior CRA history (red). The model demonstrates superior net benefit across a range of clinically relevant thresholds. The “all” (orange) and “none” (purple) lines represent the strategies of treating all or no patients, respectively.

Compared to existing literature on caries risk prediction, our study introduces several key advancements in both data infrastructure and modeling approach. First, prior studies have relied on small datasets (typically fewer than 5,000 patients), often derived from cross-sectional surveys or limited retrospective cohorts (Wang et al. 2025; Ramos-Gomez et al. 2021; Sadegh-Zadeh et al. 2022; Park et al. 2021). In contrast, our model is trained on a large-scale, longitudinal dataset curated from dental EHRs encompassing over 1 million visits across diverse sites and patient populations. Second, most of the previous studies identified predictors through traditional feature selection procedures, such as univariate or bivariate analysis followed by backward selection (Duan et al. 2021; Luo et al. 2021; Wickramasinghe and Usgodaarachchi, 2022) and multivariate analysis followed by stepwise backward selection (Reyes et al. 2023), which may introduce bias or limit generalizability. In contrast, our model used the full set of variables that have already been validated as caries-relevant and are aligned with established dental quality measures (Kumar et al. 2018; Bangar et al. 2022; Neumann et al. 2019). Our proposed approach ensures strong clinical relevance without relying on ad hoc variable screening processes seen in many previous studies (Duan et al. 2021; Luo et al. 2021; Wickramasinghe and Usgodaarachchi, 2022; Reyes et al. 2023). The updatable and longitudinal nature of EHRs offers opportunities for continuous risk monitoring and quality improvement. The use of a standardized Common Data Model (CDM) facilitates reproducible feature extraction from EHRs. While the initial data extraction currently requires manual queries, the structured nature of the CDM makes the process readily automatable. This makes our framework scalable and repeatable for ongoing risk prediction as new data becomes available in the future. Third, our predictors were mostly fed to the model after converting them into a categorical format, which helps overcome the issue of missingness and enables the training of a lower-dimensional embedding layer, which is a key element of our model. We have found that just converting our features into categorical format can improve the performance of even a simple machine learning model such as LGBM by 12% as shown in Supplementary Table 1. Fourth, while most prior work has focused on identifying static risk factors (Yan et al. 2023; Wang et al. 2025) or predicting binary caries outcomes (Park et al. 2021; Wang et al. 2025), our model takes a fundamentally different approach by estimating a personalized caries risk score using a bi-GRU time-to-event-based model. Unlike conventional binary classification models that predict only whether a patient will develop new caries within a fixed time window, our model uses a survival-based objective to estimate the timing of disease onset. This enables more nuanced, time-aware risk stratification by capturing both the likelihood and the expected timing of the outcome. From a clinical perspective, survival-based risk estimates allow providers to prioritize patients for timely preventive interventions, personalize recall schedules based on individual risk trajectories, and allocate resources more efficiently. Additionally, based on our experiments, we found our proposed survival model offers better performance within the 6 and 12-month windows compared with just using a classification-based model, as shown in Supplementary Table 3 and Supplementary Table 5.

The observed clinical benefit of our model likely stems from its ability to account for patients’ longitudinal dental histories, including the timing and sequence of visits—factors not captured by traditional models. The proposed Bi-GRU model was designed to capture patients’ longitudinal dental histories by preserving the temporal sequence of visits and the intervals between them. This approach enables the model to learn dynamic risk patterns and temporal dependencies across time, leveraging the core strength of RNN in modeling sequential data. In contrast, traditional machine learning models such as LR and LGBM rely on flattened, aggregated inputs, which omit essential temporal information such as visit ordering and spacing. Although LGBM achieved a competitive AUROC, the Bi-GRU model surpassed it by incorporating the full temporal structure of patient histories.

The subgroup analysis demonstrates the robustness of our model across a diverse patient population. The high c-index values observed across insurance types, age groups, and racial groups suggest the model generalizes well to different socioeconomic and developmental profiles. Notably, performance gains in older adolescents and patients with higher CRA levels may reflect increased signal from accumulated clinical history or elevated risk patterns that are more easily learnable by the model. Although slightly lower performance was observed in very young and cash-paying patients, these remain within an acceptable range and may reflect limited data density or clinical variability in these groups. Overall, the consistent model performance supports its applicability for risk stratification across demographic and clinical strata.

This study leveraged data extracted from EHR platforms across multiple clinical sites. Although we prioritized features known to be highly relevant for caries risk prediction, there remains considerable opportunity to enhance model inputs by incorporating a broader range of data sources across clinical domains. One key variable in our model is the CAMBRA-CRA score—a composite measure based on dental inspection, patient history, and structured questionnaires. While the CRA level is predictive, it functions as a summary metric; future work should aim to deconstruct this score and incorporate its individual components directly as model features. Doing so could improve interpretability, enhance clinical utility, and increase generalizability across patient subgroups. Additionally, the quality and consistency of provider-specified CRA assessments present a limitation, as variability in clinical judgment and documentation may introduce noise. Notably, predictive tools such as ours may help mitigate this issue by offering standardized, data-driven risk estimates to support and refine clinical decision-making.

## Conclusion

In this study, we developed and validated an RNN-based model leveraging longitudinal dental records to predict future caries risk with high accuracy and generalizability. By integrating temporal patterns and multi-domain clinical features across visits, the proposed Bi-GRU model significantly outperformed traditional machine learning models and established clinical baselines such as CAMBRA-CRA. Decision curve analysis further demonstrated the model’s superior net clinical benefit across a wide range of risk thresholds, highlighting its potential to support more precise and proactive decision-making in dental care. These findings underscore the value of harnessing structured EHR data and sequential modeling techniques to improve risk stratification and personalize preventive strategies. Future work should focus on expanding data inputs, validating the model prospectively, and integrating it into EHR systems so that risk predictions can be generated in real time during patient visits. Such integration would also allow the model to be continuously updated with new patient records and contribute to the broader vision of precision medicine, in which oral health is an integral component of whole-person health.

## Data Availability

All data produced in the present study are available upon reasonable request to the authors

## References

GBD 2021 Oral Disorders Collaborators. Trends in the global, regional, and national burden of oral conditions from 1990 to 2021: a systematic analysis for the Global Burden of Disease Study 2021. Lancet. 2025 Mar 15;405(10482):897–910. doi: 10.1016/S0140-6736(24)02811-3. Epub 2025 Feb 27. PMID: 40024264.

Wen PYF, Chen MX, Zhong YJ, Dong QQ, Wong HM. Global Burden and Inequality of Dental Caries, 1990 to 2019. J Dent Res. 2022 Apr;101(4):392–399. doi: 10.1177/00220345211056247. Epub 2021 Dec 2. PMID: 34852668.

Bratthall D, Hänsel Petersson G. 2005. Cariogram: a multifactorial risk assessment model for a multifactorial disease. Community Dent Oral Epidemiol. 33(4):256–264.

Featherstone JDB, Chaffee BW. 2018. The evidence for caries management by risk assessment (CAMBRA®). Adv Dent Res. 29(1):9–14.

Gao XL, Hsu C-YS, Xu Y, Hwarng HB, Loh T, Koh D. 2010. Building caries risk assessment models for children. J Dent Res. 89(6):637–643.

Brons-Piche E, Eckert GJ, Fontana M. Predictive Validity of a Caries Risk Assessment Model at a Dental School. J Dent Educ. 2019 Feb;83(2):144–150. doi: 10.21815/JDE.019.017. PMID: 30709989.

Paul P, Kalghatgi S, Dalvi T, Magdum SS. Advancing Dental Risk Profiling: A Literature Review of the Cariogram Model. Cureus. 2025 Mar 5;17(3):e80069. doi: 10.7759/cureus.80069. PMID: 0190924; PMCID: PMC11969287.

Lucchi P, Nasuti AD, Franciosi G, Gaeta C, Grandini S, Ludovichetti FS, Mazzoleni S, Malvicini G. 2024. Introducing the Index of Caries Risk (ICR): a comparative study on a novel tool for caries risk assessment in pediatric patients. Children. 11(10):1166.

Su N, Lagerweij MD, van der Heijden GJMG. Assessment of predictive performance of caries risk assessment models based on a systematic review and meta-analysis. J Dent. 2021 Jul;110:103664. doi: 10.1016/j.jdent.2021.103664. Epub 2021 May 10. PMID: 33984413.

Ng TCH, Luo BW, Lam WYH, Baysan A, Chu CH, Yu OY. 2024. Updates on caries risk assessment—a literature review. Dent J (Basel). 12(10):312.

Brandon RG, Bangar S, Yansane A, Neumann A, Mullins JM, Kalenderian E, et al. 2022. Development of quality measures to assess tooth decay outcomes from electronic health record data. J Public Health Dent. 83(1):33–42.

Moons KGM, Wolff RF, Riley RD, Whiting PF, Westwood M, Collins GS, Reitsma JB, Kleijnen J, Mallett S. PROBAST: A Tool to Assess Risk of Bias and Applicability of Prediction Model Studies: Explanation and Elaboration. Ann Intern Med. 2019 Jan 1;170(1):W1–W33. doi: 10.7326/M18-1377. PMID: 30596876.

Wang X, Zhang P, Lu H, Luo D, Yang D, Li K, et al. 2025. Risk prediction models for dental caries in children and adolescents: a systematic review and meta-analysis. BMJ Open. 15(3):e088253.

Ramos-Gomez F, Marcus M, Maida CA, Wang Y, Kinsler JJ, Xiong D, et al. 2021. Using a machine learning algorithm to predict the likelihood of presence of dental caries among children aged 2 to 7. Dent J (Basel). 9(12):141.

Sadegh-Zadeh SA, Rahmani Qeranqayeh A, Benkhalifa E, Dyke D, Taylor L, Bagheri M. 2022. Dental caries risk assessment in children 5 years old and under via machine learning. Dent J (Basel). 10(9):164.

Park YH, Kim SH, Choi YY. 2021. Prediction models of early childhood caries based on machine learning algorithms. Int J Environ Res Public Health. 18(16):8613.

Kang IA, Ngnamsie Njimbouom S, Lee KO, Kim JD. 2022. DCP: prediction of dental caries using machine learning in personalized medicine. Appl Sci. 12(6):3043.

Rasmy L, Xiang Y, Xie Z, et al. 2021. Med-BERT: pretrained contextualized embeddings on large-scale structured electronic health records for disease prediction. NPJ Digit Med. 4:86.

Jagannatha AN, Yu H. 2016. Bidirectional RNN for medical event detection in electronic health records. Proc Conf. 2016:473–482. doi:10.18653/v1/N16-1056.

Rasmy L, Zhu J, Li Z, et al. 2021. Simple recurrent neural networks is all we need for clinical events predictions using EHR data. arXiv. 2110.00998.

Rasmy L, Nigo M, Kannadath BS, Xie Z, Mao B, Patel K, Zhou Y, Zhang W, Ross A, et al. 2022. Recurrent neural network models (CovRNN) for predicting outcomes of patients with COVID-19 on admission to hospital: model development and validation using electronic health record data. Lancet Digit Health. 4(6):e415–25.

Nigo M, Rasmy L, Mao B, Kannadath BS, Xie Z, Zhi D. 2024. Deep learning model for personalized prediction of positive MRSA culture using time-series electronic health records. Nat Commun. 15(1):2036.

Bangar S, Neumann A, White JM, Yansane A, Johnson TR, Olson GW, et al. 2022. Caries risk documentation and prevention: eMeasures for dental electronic health records. Appl Clin Inform. 13(1):80–90.

Kumar AA, Ananthakrishnan MG, Kumar S, Divakar G, Sekar S, Dharani S. 2022. Assessing the validity and reliability of tooth widths and Bolton ratios obtained from digital models and plaster models. J Pharm Bioallied Sci. 14(Suppl 1):S148–51.

Neumann A, Obadan-Udoh E, Bangar S, Kumar SV, Tokede O, Kim A, et al. 2019. Number of pregnant women at four dental clinics and the care they received: a dental quality eMeasure evaluation. J Dent Educ. 83(10):1158–65.

Wu S, Liu S, Sohn S, Moon S, Wi CI, Juhn Y, Liu H. 2018. Modeling asynchronous event sequences with RNNs. J Biomed Inform. 83:167–77.

White JM, Brandon RG, Mullins JM, Simmons KL, Kottek AM, Mertz EA. 2020. Tracking oral health in a standardized, evidence-based, prevention-focused dental care system. J Public Health Dent. 80(S2). doi:10.1111/jphd.12413

Vickers AJ, van Calster B, Steyerberg EW. 2019. A simple, step-by-step guide to interpreting decision curve analysis. Diagn Progn Res. 3:18.

Duan S, Li M, Zhao J, Yang H, He J, Lei L, et al. 2021. A predictive nomogram: a cross-sectional study on a simple-to-use model for screening 12-year-old children for severe caries in middle schools. BMC Oral Health. 21(1):457.

Luo Y, Zhang H, Zeng X, Xu W, Wang X, Zhang Y, Wang Y. 2021. Nomogram prediction of caries risk among schoolchildren age 7 years based on a cohort study in Shanghai. J Int Med Res. 49(11):3000605211060175.

Wickramasinghe D, Usgodaarachchi U. 2022. Identifying high-risk children for dental caries in school settings: a simple predictive model. Ceylon Med J. 67(4):157–63.

Toledo Reyes L, Knorst JK, Ortiz FR, Brondani B, Emmanuelli B, Saraiva Guedes R, et al. 2023. Early childhood predictors for dental caries: a machine learning approach. J Dent Res. 102(9):999–1006.

Yan X, Sun T, Lu Y, Tan X, Wang Z, Li M. 2023. Prediction model of dental caries in 12-year-old children in Sichuan Province based on machine learning. Hua Xi Kou Qiang Yi Xue Za Zhi. 41(6):686–93.

