## Supplementary Material for "RT2C: Predicting Time to New Dental Caries Using a Recurrent Neural Network Model Trained on Multi-Site Longitudinal Structured Dental Data"

### Supplementary Appendix

### Data Preparation

For the same training cohort, we created model inputs in two formats. One treated continuous features as they are, and the other one treated continuous features as categorical while keeping their original values and not specifying any predefined discretization. Then we fit two separate light gradient boosting machine (LGBM) models with two inputs. The performance gain of treating continuous features as categorical is superior (Table 1).

*Table 1.*

| | As continuous features | As categorical features | p-value[2] |
|---|---|---|---|
| AUROC[1] (std) | 0.70 (0.12) | 0.82 (0.05) | < 0.005 |

1. Area under the receiver operating characteristic curve
2. Welch's t-test

### Data Structuring and Model Architecture

For each visit, its dental history was structured into a four-layered list as illustrated in Fig 1. The outermost layer contains the visit identifier, the binary new caries outcome label, time to event in days between the visit and the previous visit; then it follows with a list representing the dental history, where each visit was represented by a sub-list containing the duration between the visit and the previous one and feature-value pairs. After embedding, the time difference between two consecutive visits were appended to the embedded vectors. By doing so, the model improved the predictive performance by incorporating the temporal knowledge (Wu et al. 2018). The proposed Bi-GRU model consisted of two hidden states, which processed sequential data in both forward and backward directions using GRUs. The outputs from both directions were then combined to generate the final prediction.

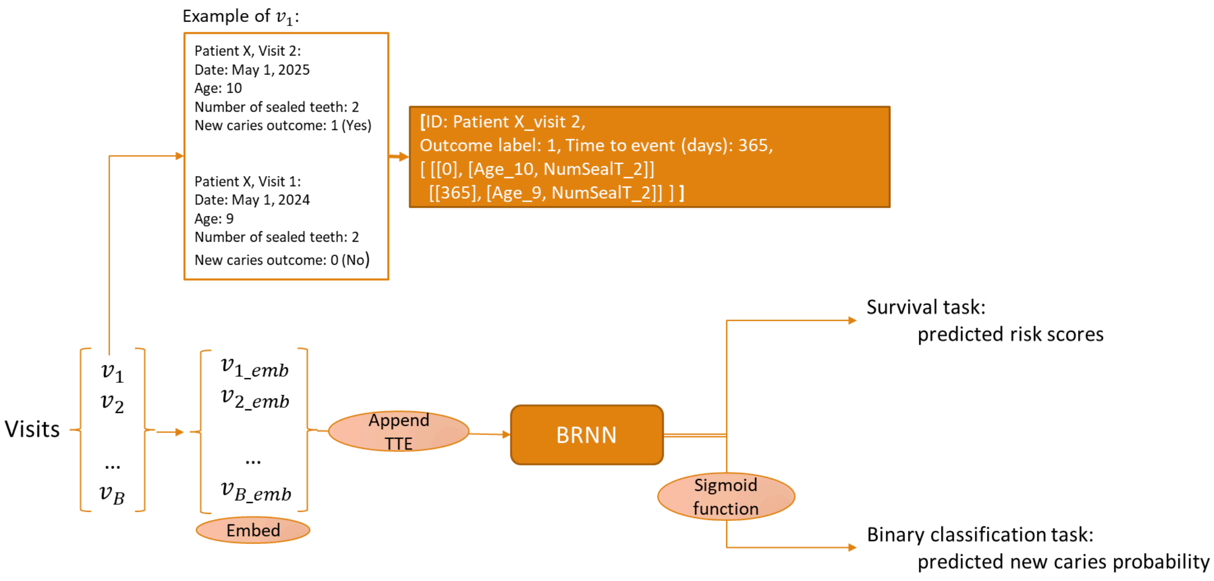


*Fig. 1* For each visit, the entire dental history is used for model prediction. For illustration purposes, only a subset of features are listed. *B*: Batch size. *TTE*: Time to event.

### Survival Prediction Performance

*Table 2*. Survival prediction performance with different cell units in RNN

| | Bi-GRU | GRU | Bi-LSTM | LSTM | p-value* |
|---|---|---|---|---|---|
| Test c-index (std) | 0.88 (0.01) | 0.87 (0.00) | 0.86 (0.02) | 0.88 (0.01) | 0.09 |

*ANOVA

*Table 3*. Performance Metrics of the Survival Model Across Timepoints

| Time window (months) | Event Rate | AUROC[1] | AUPRC[1] | Spec95[2] | Sensitivity | Specificity | F1-score |
|---|---|---|---|---|---|---|---|
| 2 | 0.00 | 0.66 | 0.00 | 0.60 | 1.00 | 0.19 | 0.00 |
| 3 | 0.00 | 0.76 | 0.00 | 0.40 | 1.00 | 0.19 | 0.00 |
| 6 | 0.00 | 0.88 | 0.08 | 0.54 | 1.00 | 0.19 | 0.01 |
| 12 | 0.06 | 0.87 | 0.26 | 0.59 | 1.00 | 0.20 | 0.13 |
| 16 | 0.13 | 0.77 | 0.26 | 0.45 | 1.00 | 0.22 | 0.27 |
| 24 | 0.17 | 0.67 | 0.26 | 0.20 | 0.94 | 0.22 | 0.33 |
| 36 | 0.21 | 0.62 | 0.27 | 0.13 | 0.89 | 0.21 | 0.36 |

| | | | | | | | |
|---|---|---|---|---|---|---|---|
| 72 | 0.22 | 0.59 | 0.27 | 0.10 | 0.87 | 0.21 | 0.37 |

1. Calculated based on the thresholding probability that maximizes the metric.
2. Specificity reported at sensitivity ≥ 0.95.

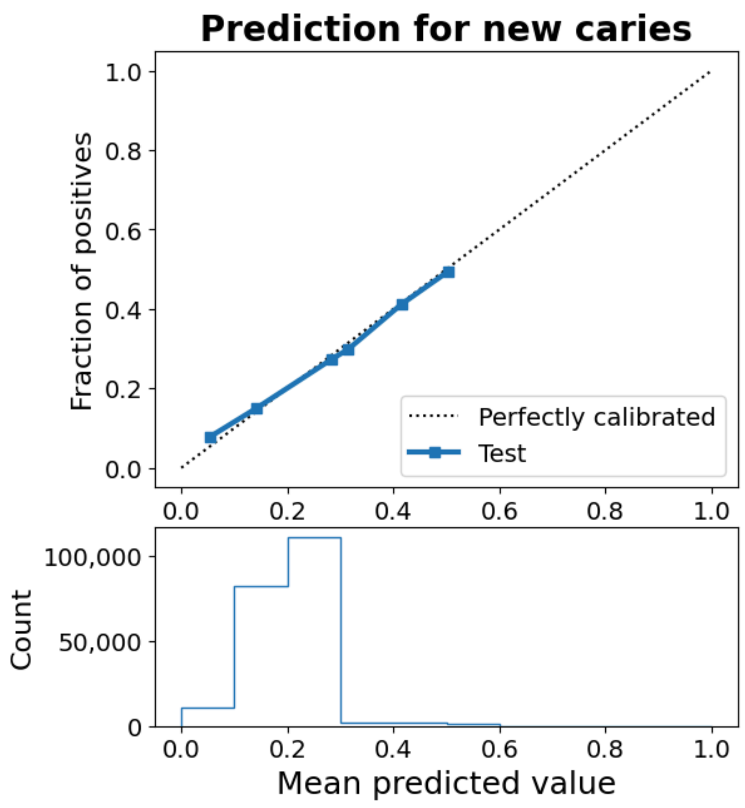


*Fig 3.* Calibration on model predicted probability of new caries at next visit

The model-predicted event probabilities were well-calibrated as the calibrated probabilities were aligned with the actual outcomes (Fig. 3). After calibration, the model's predictions are concentrated in the 0.1–0.3 range, with few predictions above 0.5. The result was consistent with the actual event prevalence which is around 0.2.

**Binary Prediction Performance**

In addition to the survival prediction task, the proposed model for binary prediction task of new caries at next visit achieved AUROC of 82.59% (95% confidence interval (CI): 82.57-82.61), outperforming traditional machine learning algorithms: logistic regression ($AUROC^{LR}$: 78.09% (95% CI: 78.06 - 78.11)) and light gradient boosting machine ($AUROC^{LGBM}$: 81.60% (95% CI: 81.58-81.62)).

*Table 5.* Binary Prediction of New Caries

| Evaluating Metrics | Mean (std) |
|---|---|
| AUROC | 0.83 (0.000) |
| AUPRC[1] | 0.57 (0.000) |
| Spec95[2] | 0.37 (0.004) |
| F1-score | 0.57 (0.002) |
| Optimum sensitivity[3] | 0.75 (0.003) |
| Optimum specificity[3] | 0.75 (0.004) |

[1]Area under the precision-recall curve

[2]Specificity at 95% sensitivity

[3]Sensitivity and specificity at the optimum threshold (the cutoff probability which leads to the highest sensitivity and specificity)

Given the data was highly imbalanced, where the prevalence of events was 20% across training, validation and test sets, we also evaluated the model based on area under the precision-recall curve (AUPRC). The proposed model achieved AUPRC of 57.71% (std: 0.28), which was substantially higher than the event prevalence rate of 20%. Other clinically relevant metrics are reported in Table 5. Fig.3 is the calibration plot with y-axis showing the actual proportion of positive outcomes, i.e., having new tooth decay, for visits that fall into the given predicted probability bin. Moreover, we compared AUROC above to AUROC achieved by employing measures from the most recent visit (82.33%, 95%CI: 82.28-82.38%) and found no significant difference.

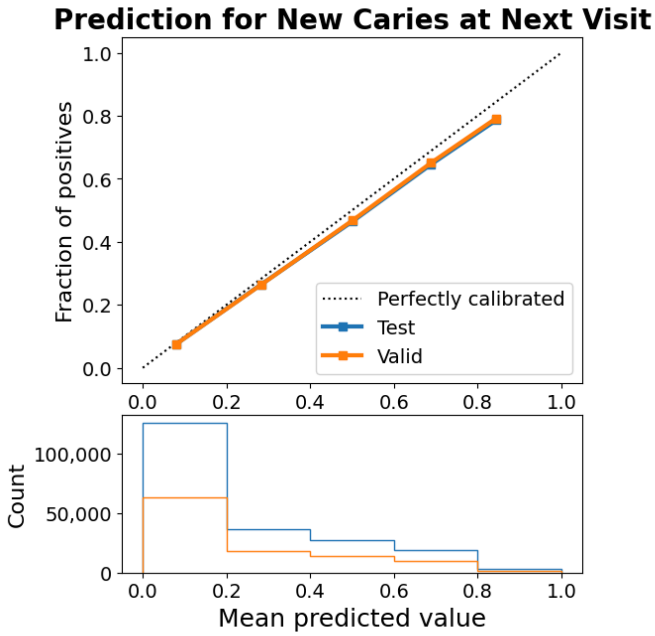


Fig 2. Calibration plot of binary prediction.
